# Life course shaping of allostatic load across 17 countries: Evidence from England, the U.S., Europe, China and Indonesia

**DOI:** 10.64898/2026.07.29.26359214

**Authors:** Guannan Li, Vanessa De Rubeis, Matteo Cesari, Ritu Sadana, Chandni Maria Jacob, Hsin-yi Lee, Gindo Tampubolon

## Abstract

Early life adversity is a recognised risk factor for poor health in later life, but its relationship with allostatic load (AL; a measure of cumulative physiological stress) remains underexplored across populations around the world. This study builds evidence to test a life course approach to AL using harmonised data from six longitudinal ageing cohorts: Health and Retirement Study (US, N=2,427), English Longitudinal Study of Ageing (England, N=2,038), The Irish Longitudinal Study on Ageing (Ireland, N=8,184), Survey of Health Ageing and Retirement in Europe (Europe, N=4,079), China Health and Retirement Longitudinal Study (China, N=5,220), and Indonesia Family Life Survey (Indonesia, N=1,243). Childhood information at aged ten to sixteen was collected retrospectively from adults aged 65 on average. An AL score is constructed using biomarker data and a latent construct of early life adversity constructed to address recall bias. Results show that early life poverty is significantly associated with raised AL in Ireland, China, and Indonesia, supporting the impact of early life experiences on older adulthood. However, studies from other countries do not confirm statistical significance. These findings document that experiencing poverty during development phase in some settings is associated with higher AL in later life, however further analyses are required to explore possible variations across different populations and their social and economic context. These results add further evidence that social determinants of health, including child poverty, in some settings, contribute to cumulative physiological stress across the life course, and that policies and actions to reduce child poverty can have beneficial effects not only as children, but also as older adults.

## Introduction

Virginia Woolf observed nearly a century ago ‘On being ill’ [1],

> People write always on the doings of the mind, the thoughts that come into it, its noble plans, how the mind has civilised the universe. They show it ignoring the body in the philosopher’s turret [But] The creature within can only gaze from the pane – smudge or rosy; it cannot separate off from the body like the pod of a pea; it must go through the whole unending process of changes, heat and cold, comfort and discomfort, hunger and satisfaction, health and illness, until there comes the inevitable catastrophe; the body smashes itself to smithereens, and the soul escapes. But of all this daily drama of the body there is no record.

Of course, there is: the faithful record is called allostatic load. Allostatic load (AL) is measured by an index that quantifies cumulative physiological wear and tear resulting from chronic stress exposures throughout the life course. Early life adversity or childhood poverty – such as heat, cold, discomfort or hunger that are markers of the experience of poverty – is a well-established risk factor for disability, dysfunction, and disease in later life across diverse contexts and countries [2–12]. But how early life adversity as a social and economic determinant of health impacts underlying biological mechanisms that trigger subsequent disability and or worsening of a person’s health trajectory, is important to establish. Evidence on plausible biological mechanisms not only strengthens the pathway between these broader determinants and outcomes, but importantly, could provide insight on what types of interventions could mitigate the negative impact of root or distal determinants of health. On one hand, AL is documented as a robust longitudinal predictor of self-rated health, physical functioning and mortality in the US and the UK [13, 14]. It also shows clear socioeconomic gradients, mirroring disparities in income, education, and occupational status [14]. As a multisystemic marker, AL can capture the risk profile of an individual for major adverse clinical outcomes, higher healthcare costs, and mortality [15, 16].

On the other, drawing on life course epidemiology and epigenetic modifications [4, 17–19], a growing body of research suggests that early life adversity particularly the experience of poverty, characterised by material deprivation and psychosocial stress, can provoke prolonged activation of the hypothalamic-pituitary-adrenal (HPA) axis and the sympathetic nervous system. Such chronic physiological arousal is theorised to disrupt regulatory functions across multiple biological systems, ultimately contributing to physiological dysregulation in adulthood. It is often measured as AL [20, 21]. Yet there are no studies that look at the impact of early life adversity on AL directly across the life course. This is partly due to the scarcity of harmonised biomarkers and retrospective childhood data in development and ageing studies.

This paper therefore addresses an important research gap and investigates the association between early life poverty on AL measured in later life, drawing on existing nationally representative studies, in a wide range of countries. Moreover, by examining whether this association exists in a wide range of countries including high- and middle-income, this study contributes to generalising the impact of broader determinants of health across the life course.

## Methods and materials

### Study design and data sources

Data from six ageing cohort studies (HRS-family) are used: Health and Retirement Study (HRS) [22], English Longitudinal Study of Ageing (ELSA) [23], Survey of Health Ageing and Retirement in Europe (SHARE, 12 countries with blood biomarkers) [24], The Irish Longitudinal Study on Ageing (TILDA) [25], China Health and Retirement Longitudinal Study (CHARLS) [26] and Indonesia Family Life Survey (IFLS) [27]. To maximise the analytic sample, this study merged early life and biomarkers information, ensuring the contemporaneous or most recent biomarkers for each survey: HRS (wave 11, 2016), ELSA (wave 8, 2016), TILDA (wave 1, 2009–2011), SHARE (wave 6, 2015), CHARLS (wave 1, 2011), and IFLS (wave 5, 2015). Childhood information was collected at various years: HRS in 2015, 2017, 2019; ELSA in 2006; CHARLS in 2014; SHARE in 2008-2009; TILDA in 2009; IFLS in 2015. We included all survey participants as described shortly. Together, these studies cover 35% of the global population and 51% of those aged 50 and over, offering new global evidence in a field where 90% of articles in leading ageing journals focus on high-income countries, with most of the ageing population living elsewhere [28]. See S1 Table, S2 Table, and S3 Table for biomarker coverage, harmonised childhood items, and selection patterns.

### Outcome: Allostatic load

Per prior practice which made a virtue of available data, we included different numbers of biomarkers in HRS, ELSA, TILDA, SHARE, CHARLS and IFLS. See S1 Table. Consistent with previous studies [29–33], we calculated AL scores for each of the HRS, ELSA, TILDA, SHARE, CHARLS, IFLS in three steps. First, we excluded respondents with missing data on any of the AL components and, following prior practice, we checked the characters of the excluded and remaining/analytic samples [10]. Second, we standardised each biomarker to have a Z score. Third, we take the mean of the Z scores, giving one new AL score that can be interpreted in terms of standard deviation units. A score greater than one standard deviation above the mean would place an adult in approximately the top 16% for physiological dysregulation. Put differently, the further the score extends beyond one standard deviation, the greater the proportion of adults experiencing elevated physiological dysregulation. Higher values indicate greater multisystem physiological dysregulation [29]. Analytic weights were applied in each survey where weights were available.

### Key exposure: Childhood poverty

The participants were asked retrospectively about their childhood –in some cases reflecting on events eight decades earlier. Obtaining information on childhood adversity from adults with an average age of 65 year raises concern about recall errors. For example, in 2008 some 2500 Britons aged 50 were asked to recall the numbers of rooms and people in their homes when they were eleven to assess overcrowding (the British 1958 cohort) [2, 3, 34, 35]. Two thirds got the numbers wrong [35]. Their mothers gave the right answers when visited four decades earlier. Thus, it is scarcely plausible that an average 65-year-old’s recall is free of errors. As an illustration, Tampubolon [3] found that, “material poverty when growing up shows no association with health when growing old, assuming accurate recall. Once recall problems are controlled, [it is] found that childhood material poverty changes inversely with later life health.” See Brown [36] for another illustration. Assuming error-free retrospective information is unsafe.

Following prior practice using latent constructs to address recall error, we constructed a binary latent class of childhood poverty (poor or non-poor) [2–4, 10–12]. More important than unsafe inference, where ignoring recall problems led to erroneous associations, using retrospective information unprocessed also prevents cross-country learning. For example, to explain the health of older Chinese both Si and colleagues and Li and colleagues used retrospective childhood conditions collected in CHARLS including death from starvation during China’s Great Leap Forward [9, 37]. But that information, unprocessed, complicates comparison with other countries’ childhood conditions because no government in the U.S., England, Europe, Ireland and Indonesia ever imposed a comparable edict. Conversely, an indicator of the presence of running hot water in ELSA and SHARE will equally complicate the comparison because this was not asked in CHARLS, HRS, TILDA or IFLS. Without harmonisation, these variations complicate comparative analysis. To overcome this, researchers from America, England, Europe and China have devised a latent construct for early life adversity or childhood poverty (harmonised) which enables it to be used as a risk factor to explain health in later life [2–6, 8, 10–12]. The early life indicators in HRS, CHARLS, ELSA, SHARE and IFLS we used here have been described elsewhere [3, 4, 10–12, 19]. See S2 Table.

### Covariates

Previous literature on characteristics associated with AL in later life was used to determine potential covariates including age, sex, marital status (unmarried/divorced/separated/widowed, married/union), residence (rural vs. urban, living in urban or living in suburban/ex-urban is used in HRS, living in Dublin or not is used in TILDA, hukou or registered as rural resident is used in CHARLS), wealth quartiles and depression. Education is coded as a threefold ordinal variable: up to primary education, secondary and college/more [30, 31].

### Analysis

Descriptive statistics were calculated for each study (means and standard deviations for continuous variables; frequencies and percentages for categorical ones). As AL is constructed as a Z score average, we fitted six linear regressions, one for each survey or cohort. We adjusted standard errors and confidence intervals for the use of derived early life adversity as the exposure following prior practice [2–4, 10, 11, 38]. Missing data were tested using chi-square for binary variable and t tests for continuous variable. Significance level is set at 5 percent. To ease interpretation, we produced marginal plots of predicted AL scores across age 50 to 90 year, distinguishing the childhood poor from the non-poor. Estimation is done in Latent GOLD version 6.1 and plotting is done in Stata version 19.

## Results

### Missing data and selection for retrospective interviews

We begin by describing the analytic sample which comprises prospective sample members who gave complete AL biomarker measurements and who agreed to retrospective interviews. Not all of them did and there may be significant differences between the analytic vs declined sample See S3 Table. Compared to those with missing data, our analytic sample does not differ by sex and age in HRS. Also, there is no sex difference in ELSA or CHARLS. However, we observe age differences in ELSA, SHARE, TILDA, CHARLS and IFLS. In ELSA, SHARE, and CHARLS, participants in the analytic sample tend to be older, whereas in TILDA and IFLS, the mean age is younger. There is no systematic pattern in the composition of the analytic samples. This lack of systematic patterns has been observed in the previous study on the life course shaping of frailty [10].

Table 1 summarises the analytic sample for each country. Among high income countries, the US has the lowest rate of childhood poverty (12.3%) and Ireland has the highest (20.1%). Among older adults in the middle income countries, China has 9.7% and Indonesia has 13.1% rates of childhood poverty. There were slightly more women in the sample compared to men in all regions, ranging from 51.6% in China to 61.7% in Indonesia. The US has the highest proportion of older adults with some college education (58.3%), and China the lowest (0.9%). In the US, 31.6% of respondents reported being divorced, separated, widowed or never married compared to 3.1% in China. On financial assets, 23.1% of older Americans fall into the poorest quartile, while Europe reports the lowest.

**Table 1.**
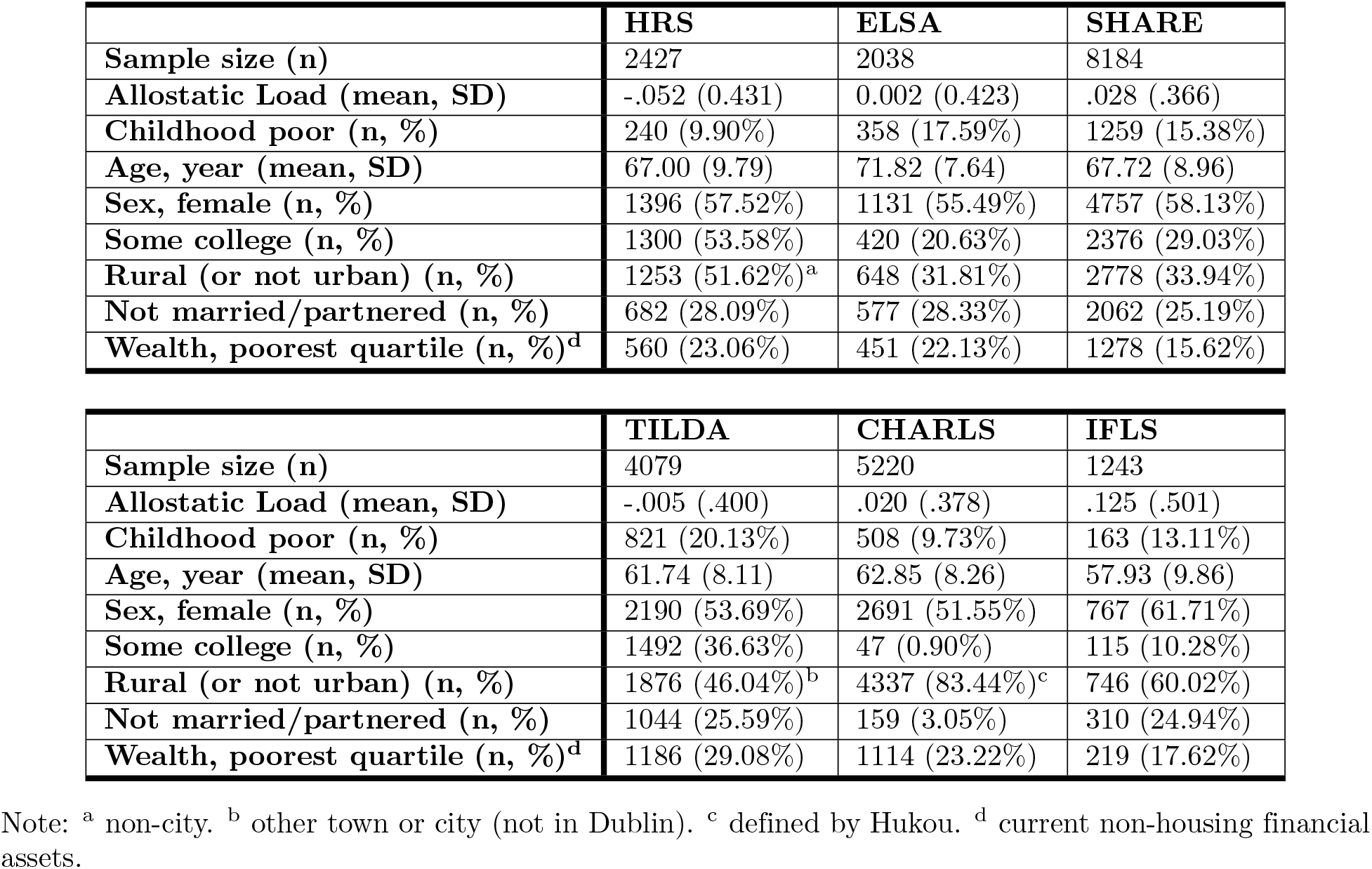
Descriptive statistics of the analytic samples.

|  | HRS | ELSA | SHARE |
| --- | --- | --- | --- |
| Sample size (n) | 2427 | 2038 | 8184 |
| Allostatic Load (mean, SD) | -.052 (0.431) | 0.002 (0.423) | .028 (.366) |
| Childhood poor (n, %) | 240 (9.90%) | 358 (17.59%) | 1259 (15.38%) |
| Age, year (mean, SD) | 67.00 (9.79) | 71.82 (7.64) | 67.72 (8.96) |
| Sex, female (n, %) | 1396 (57.52%) | 1131 (55.49%) | 4757 (58.13%) |
| Some college (n, %) | 1300 (53.58%) | 420 (20.63%) | 2376 (29.03%) |
| Rural (or not urban) (n, %) | 1253 (51.62%)<sup>a</sup> | 648 (31.81%) | 2778 (33.94%) |
| Not married/partnered (n, %) | 682 (28.09%) | 577 (28.33%) | 2062 (25.19%) |
| Wealth, poorest quartile (n, %)<sup>d</sup> | 560 (23.06%) | 451 (22.13%) | 1278 (15.62%) |

|  | TILDA | CHARLS | IFLS |
| --- | --- | --- | --- |
| Sample size (n) | 4079 | 5220 | 1243 |
| Allostatic Load (mean, SD) | -.005 (.400) | .020 (.378) | .125 (.501) |
| Childhood poor (n, %) | 821 (20.13%) | 508 (9.73%) | 163 (13.11%) |
| Age, year (mean, SD) | 61.74 (8.11) | 62.85 (8.26) | 57.93 (9.86) |
| Sex, female (n, %) | 2190 (53.69%) | 2691 (51.55%) | 767 (61.71%) |
| Some college (n, %) | 1492 (36.63%) | 47 (0.90%) | 115 (10.28%) |
| Rural (or not urban) (n, %) | 1876 (46.04%)<sup>b</sup> | 4337 (83.44%)<sup>c</sup> | 746 (60.02%) |
| Not married/partnered (n, %) | 1044 (25.59%) | 159 (3.05%) | 310 (24.94%) |
| Wealth, poorest quartile (n, %)<sup>d</sup> | 1186 (29.08%) | 1114 (23.22%) | 219 (17.62%) |

Note: <sup>a</sup> non-city. <sup>b</sup> other town or city (not in Dublin). <sup>c</sup> defined by Hukou. <sup>d</sup> current non-housing financial assets.

Fig 1 presents smoothed empirical densities of AL for each cohort, distinguishing the childhood poor in dashed line from the non-poor in solid line. First, comparing the overall shape of densities shows that high income nations have narrower ranges of AL (e.g. ELSA); conversely, middle income nations have fatter right tails (more adults are found beyond the one unit on the horizontal axis) suggesting more adults have high physiological loads in these nations (e.g. CHARLS or IFLS).

**Fig 1.**
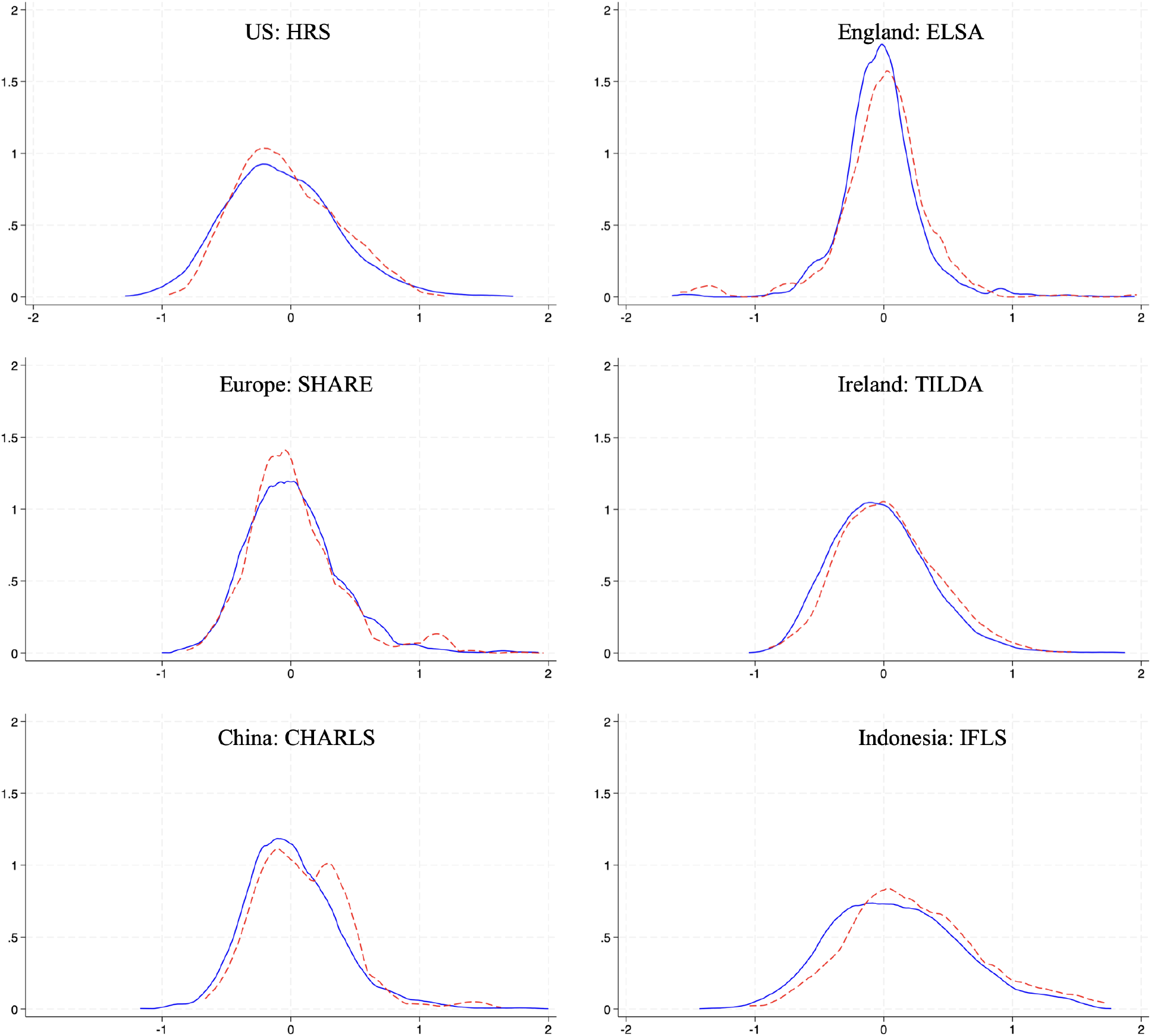
Smoothed densities of AL in HRS, ELSA, SHARE, TILDA, CHARLS, IFLS. Distinguishing the childhood poor in red dashed lines from the non-poor in blue solid lines.

More importantly, the childhood poor densities –in dashed red lines– are shifted rightward in China and Indonesia compared to in England. Put differently, it is easier to peel away the dashed red lines in China and Indonesia than in England. But the shift is not the only discernible pattern, spreading or widening is equally obvious – compare England and Indonesia again. Altogether, these shifts and widening accord with what can be expected given the average incomes of these countries, thus offering some informal external validity. In sum, middle income nations have proportionately more adults with high AL than high income nations.

Furthermore, the childhood poor in middle income nations also have disproportionately more adults with high AL.

Table 2 shows the association between early life adversity and AL for each cohort. There is no significant association between early life adversity and later life AL in high income countries except in Ireland 0.04 (95% confidence interval 0.01 - 0.07). where it was characterized by high unemployment and mass emigration until the mid-1980s. In contrast, in China and Indonesia early adversity is significantly associated with higher AL. The patterns first noted in the univariate smoothed densities remained after adjustment for covariates in the regression models, suggesting that irrespective of sex, age and other life course attainments such as education, there is a significant shift to higher AL among those whose childhood was poor. To ease interpretation, we shall plot marginal or predicted AL for each cohort, distinguishing the childhood poor from the non-poor later. (See S4 Table for the complete results where simpler models with age and sex were fitted for each cohort.)

**Table 2.** Linear regression coefficients and confidence interval (95%) of allostatic load in the US (HRS), England (ELSA), Europe (SHARE), Ireland (TILDA), China (CHARLS) and Indonesia (IFLS)

|  | HRS | ELSA | SHARE |
| --- | --- | --- | --- |
| N. Obs | 2427 | 2038 | 8184 |
| Childhood poor | .037 (-.039, .113) | .027 (-0.018, 0.072) | .007 (-.014, .028) |
| Age | .005 (.003, .006) | .003 (.001, .005) | .002 (.001, .003) |
| Sex, female | -.066 (-.101, -.032)*** | .024 (-.012, .060) | -.152 (-.168, -.136)*** |
| Some college | -.093 (-.165, -.021)* | -.050 (-.113, .012) | .010 (-.013, .033) |
| Rural (non-urban) | -.002 (-.036, .032) | -.015 (-.055, .026) | -.009 (-.025, .008) |
| Unmarried/Single | .014 (-.025, .052) | .023 (-.758, .804) | -.020 (-.038, -.001)* |
| Poorest quartile<sup>a</sup> | .198 (.149, .247)*** | .128 (.072, .183)*** | .043 (.018, .067)** |
| Constant | -.071 (-.088, -.053)*** | -.003 (-.023, .017) | .022 (.013, .0308)*** |

|  | TILDA | CHARLS | IFLS |
| --- | --- | --- | --- |
| N. Obs | 4079 | 5220 | 1243 |
| Childhood poor | .071 (.040, .101)*** | .086 (.053, .118)*** | .132 (.053, .211)** |
| Age | .011 (.010, .013) | .008 (.006, .009) | .001 (-.002, .005) |
| Sex, female | -.125 (-.149, -.101)*** | -.028 (-.049, -.008)** | -.124 (-.181, -.067)*** |
| Some college | -.060 (-.094, -.026)** | .138 (.060, .216)** | .136 (.029, .243)* |
| Rural (non-urban) | .000 (-.024, .024) | -.056 (-.082, -.029)*** | .076 (.013, .138)* |
| Unmarried/Single | .004 (-.024, .032) | .013 (-.060, .087) | .002 (-.077, .082) |
| Poorest quartile<sup>a</sup> | .032 (.001, .064)* | .023 (-.009, .054) | -.022 (-.102, .059) |
| Constant | -.020 (-.033, -.006)** | .019 (.008, .030)** | .110 (.080, .142)*** |

Note: Significant level: \*5%, \*\*1%, \*\*\*0.1%. <sup>a</sup> current non-housing financial asset. To streamline, only the primary full adjusted models are shown per cohort representing coefficients and 95% CIs.

The results further indicate that age is a significant risk factor for the accumulation of AL, while being female is associated with lower AL in later life. Education functions as a protective factor in developed countries such as the US, England, and Ireland, where it is consistently linked to lower AL. In contrast, education appears to act as a risk factor in middle income nation contexts such as China and Indonesia. Rural residence is positively associated with higher AL in China, Indonesia, and parts of Europe, highlighting spatial disparities within these regions; this association is not observed in the US and Ireland. Marital status does not exhibit a significant relationship with AL. Living in the poorest wealth quartile is consistently associated with higher AL in high income nations, but this is not observed in middle income nations. These findings emphasise a disparity between high and middle income nations, which is examined in greater detail later.

The results for the middle income nations can be elaborated further. Take the two coefficients of childhood poverty and [being in the] poorest quartile. Recall that childhood poverty happened six or more decades ago whereas poorest quartile is today. In other words, poorest quartile is not a measure of poverty in childhood. Of course, childhood poverty six decades ago may be related to [being in the] poorest quartile today as posited in the life course model, specifically the pathway model of Kuh & Ben-Shlomo [39]. In the pathway model, the coefficient of childhood poverty is the direct association between childhood poverty and later life AL (accounting for being in the poorest quartile today, age, sex, education, residence, marital status which lie along the life course).

To ease interpretation, we present predicted AL scores in each cohort across age 50–90 years, distinguishing childhood poverty status into the childhood poor in dashed red lines and the non-poor in solid blue lines (these plots are known as marginal plots; Fig 2). The plots show higher ranges of AL in middle income nations than in high income nations, consistent with the smoothed densities in Fig 1. This is a novel finding that reflects worldwide socioeconomic development of these countries – as we move from the US through Ireland to Indonesia the range of AL also moves up, showing that as national average income decreases AL range increases.

**Fig 2.**
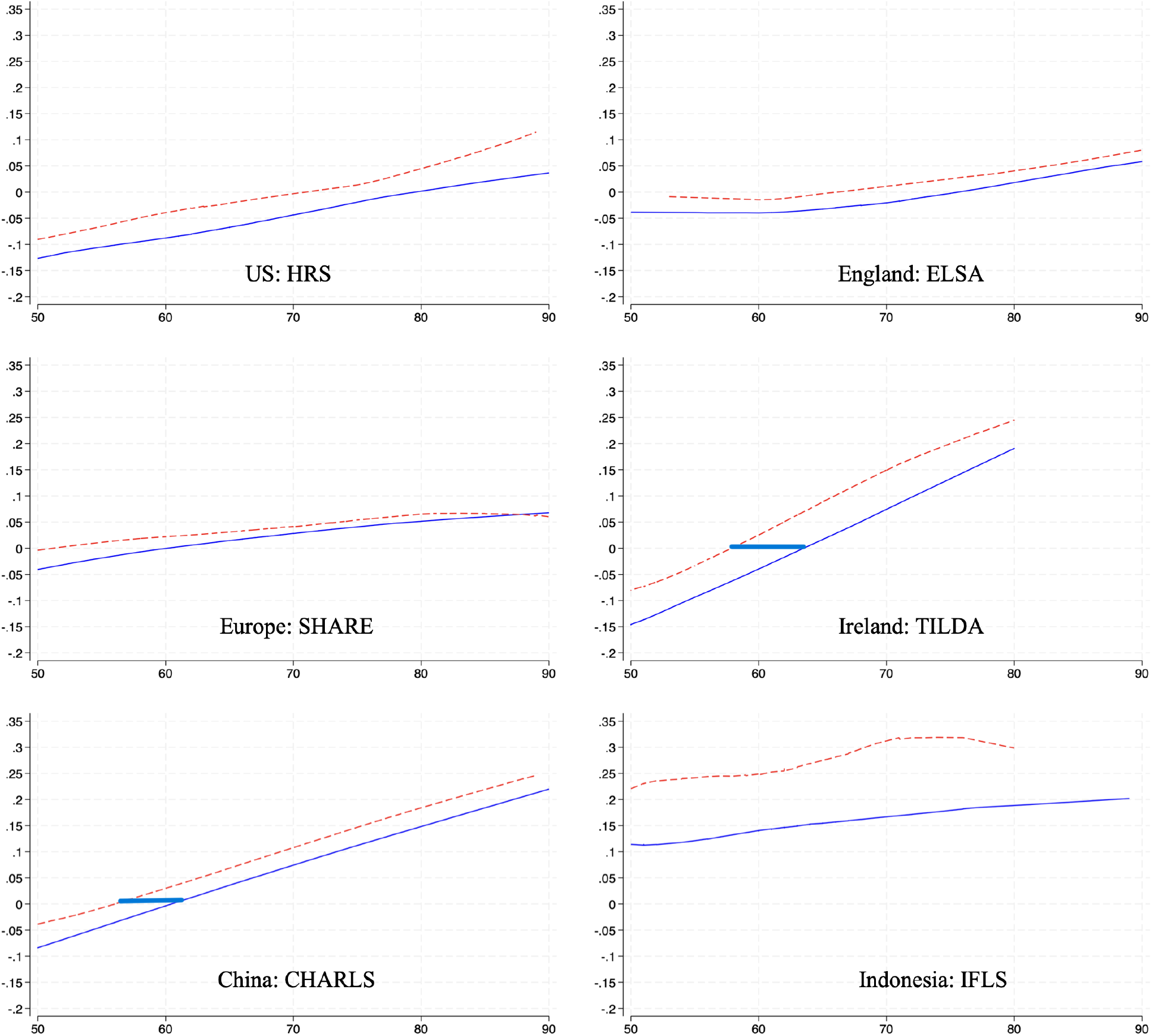
Predicted allostatic load by age and childhood poverty status across cohorts (HRS, ELSA, SHARE, TILDA, CHARLS, IFLS). The childhood poor in dashed red lines, the non-poor in solid blue lines; common age range (x-axis) and common AL range (y-axis). Distance at average AL in Ireland is 5.8 years and in China 4.3 years.

The plots demonstrate a clear upward trend in AL with advancing age, consistent with the concept of cumulative wear and tear (Fig 2). Remarkably, across all countries with wide variations in socioeconomic development, healthcare systems, and geographic and cultural contexts, adults with a history of early adversity consistently carry higher AL scores – the dashed lines are always above the solid lines. For example, take IFLS (bottom right plot): the red line is above the blue line throughout; similar to TILDA (the plot right above it): the red line is also above the blue line throughout. This pattern is repeated across all six plots or cohorts. This pattern stresses the enduring physiological burden associated with early life adversity.

Equally remarkable is the fact that the plots combined with the statistical significance in Table 2 show that in high income nations the influence of early life adversity on AL may not remain. Although the trends between childhood adversity versus no adversity remains distinct, the gap between these trends lessens among high income countries (Table 2). There is no significant difference in AL among those who had earlier adversity compared to those who had no earlier adversity (Fig 2). In sum, not only did AL accumulate higher in middle income nations (last two plots i.e. for China and Indonesia), AL in later life remains shaped by early life adversity. In contrast, the range of AL decreases and narrows as nations progresses along socioeconomic development (to join the high-income nations group). Moreover, in high income nations early life adversity also ceases to shape AL in later life. This has never been reported before.

To help interpret the significant associations with childhood poverty, following previous scheme [2], we draw the horizontal distance between the childhood poor and non-poor at average AL (zero). It shows that in Ireland the childhood poor aged 5.8 years older than the non-poor; in China the comparable figure is 4.3 years. Indonesia has the other significant childhood poverty association, but a similar exercise would lead to an unwarranted extrapolation to an earlier age. Furthermore, the other cohorts (from high income nations) have non-significant associations hence no distances are drawn.

## Discussion

If in Woolf’s essay the body records faithfully, in this study the mind recalls childhood unreliably [1–3]. Once this is addressed, the life course approach suggests that early life adversity exerts a lasting influence on later life physiological dysregulation regardless of adult protective factors including education and wealth. With recent collections of blood biomarkers, this study presents for the first time new evidence in support of the life course approach to understanding factors that influence AL from around the world [40]. We find that early adversity is associated with higher AL in later life in Ireland, China, and Indonesia. These findings suggest that early life may be a sensitive period that has lasting effects on AL, and this remains across a range of countries at different levels of socioeconomic development [2, 3, 10, 11, 39].

Childhood poverty contributes to raised AL in later life through interrelated physiological, psychological, and socio-economic pathways. Physiologically, it triggers chronic activation of the hypothalamic-pituitary-adrenal axis and sympathetic nervous system, resulting in prolonged exposure to stress hormones (e.g. cortisol), immune dysregulation, and cardiometabolic risk. Psychologically, adverse early environments increase perceived stress, emotional dysregulation, and mental health problems, which disrupt stress-response systems and coping behaviours [41, 42]. Unfortunately, only ELSA has collected cortisol level for some of its participants, obviating a direct examination of this path. Socially, early adversity often leads to lower educational attainment, reduced upward mobility, and chronic exposure to stressors such as poor housing and unsafe environments – factors consistently linked to raised AL [43].

Second, our findings highlight a clear disparity between high and middle income countries in the life course shaping of AL. In the US, England and Ireland, having some college education appears to protect against higher AL in later life, whereas in China and Indonesia, it is associated with increased AL. A similar contrast emerges in relation to wealth: while individuals in the lowest wealth quartile in developed countries show higher AL, such association is not significant in China and Indonesia. These contrasting findings align with previous longitudinal studies on AL conducted in both Western [30] and Asian contexts [31].

Third, our analysis shows that predicted AL increases with age and consistently indicates that adults who experienced poverty or adversity in early life face a disadvantage in AL later in life, showing elevated physiological wear and tear. Our findings demonstrate that early life adversity is associated with greater physiological dysregulation in later life. This is of concern given higher allostatic load is associated with decreased health span leading to early mortality [15, 16].

There is an emerging interest to explore the life course shaping of AL in high income nations though our study is the first to explore a broader context using data from 17 countries across high and middle income nations, giving a robust comparison. In two separate studies on Ireland and Britain, McLoughlin and colleagues found that out of eleven indicators of early life adversity, three are significantly associated with AL in Ireland while Barboza Solis and colleagues found no direct evidence of adverse childhood experience impacting on AL in Britons aged 44 using the British 1958 cohort [44, 45]. This is the same British 1958 cohort which we have studied to discover the extent of error-laced retrospective information [2, 3, 34]. Notably these studies on Ireland and Britain used different constructs of early life adversity, making fragile any attempt to compare and draw conclusions from them. Our study has harmonised early life adversity and AL with some success as evinced by the external validity of the smoothed densities of AL (Fig 1).

### Rectifying the imbalance on evidence and experience

By pooling six ongoing longitudinal ageing cohorts or surveys from high and middle income nations we also rectify the imbalance between evidence on ageing and the experience of ageing. We recently examined three high ranking journals on ageing on both sides of the Atlantic including Age and Ageing, Journals of Gerontology – series B, and the Journal of the American Geriatric Society. Out of 713 research articles, ninety percent focus solely on high income nations, six percent on both high and middle income nations, and only four percent on middle income nations [28]. (Any other top three journals are not likely to change these percentages). Our work here redresses the imbalance. But this goes beyond redressing a conspicuous imbalance, it is also good science. It builds on what has been scattered before, say on Ireland or Britain, evidence on which is encompassed in Fig 2. But our evidence extends theirs to show that the pattern accords with socioeconomic development of the countries in the sample (see the shift and spread in the smoothed densities in Fig 1). This new evidence redresses the imbalance while strengthening the life course framework around the world [40].

### Contributions to the literature

Early life could be a sensitive period when harmful exposures can have long lasting effects, and this work illuminates that possibility [39]. This contribution also extends the inquiry of the long arm of childhood conditions, a related thesis introduced by Hayward and Gorman in their study on the long-term impact of childhood poverty on midlife mortality among American males. While studies using longitudinal ageing cohorts accumulate evidence of persistent effects of early adversity on physical function, cognitive function, mental health, multimorbidity and healthy ageing [10, 11, 19, 37, 46], fewer have directly examined the association between early adversity and later AL, particularly using cross-national data along different levels of socioeconomic development.

Emerging evidence from large-scale ageing surveys across 29 trans-Atlantic countries – from the United States and Britain to Israel – supports the enduring influence of early life adversity or poverty on a range of later life outcomes, including decreased muscle strength, depression, cognitive decline, frailty, multimorbidity and healthy ageing [10–12, 19, 37, 46]. Such cross-country findings reinforce the importance of adopting a life course approach to understand population ageing, informing global initiatives and national strategies for all life stages under the United Nations Decade of Healthy Ageing [47] and the WHO Framework to implement a life course approach in practice [40].

### Strengths and limitations

This study has strengths and limitations and noting them is important for the ongoing cross-country ageing studies and their harmonisation. A key strength of this study is its life-course design, which leverages the clear temporal ordering whereby childhood poverty precedes allostatic load by several decades, strengthening inference about long-term physiological link. By harmonising early-life exposures and biomarker outcomes across longitudinal cohorts from high- and middle-income countries, this study moves beyond the largely cross-sectional or single-country literature to provide novel global insight into how childhood poverty shapes later-life health under contrasting socioeconomic contexts. Second, it is unusual in its breadth of country coverage, encompassing nations that represent about 35% of the global population and over half of the world’s older population aged 50 and over [48]. Last, the same method is replicable, easily deployed when biomarkers and life-course survey data are available for other countries, for instance the majority of countries in SHARE.

There are several limitations. The cross-sectional design of this study poses some of them. Most notably, it is difficult to establish causal relationships between childhood poverty and AL in later life. A moment’s thought would suggest that a randomised experiment (a strong design for establishing causality) may make it impossible to draw causal inference, as follows. There is a necessary long period separating the exposure (early life) and the outcome (later life). Half a century is typical and that period is long enough for participants to find out about their assignments at randomisation and, accordingly, modify their behaviours. Observational studies like this do not allow strong causal inference, although the support for the life course framework can be strengthened as demonstrated here, namely by going across countries. Furthermore, retrospective information about early life may introduce bias. But we have applied a recent advanced method (latent construct) to address this [2, 3, 10–12]. Additionally, selective survival bias may distort associations; individuals who experienced severe physiological dysregulation may not have survived into older age and thus are excluded from the sample, potentially underestimating the true effect of childhood poverty [35]. Furthermore, cross-sectional data limit the capacity to adjust for unmeasured or time-varying confounders, affecting confidence in the robustness of observed associations. Lastly, because AL is measured at a single point in time, this design fails to capture the dynamic nature of physiological regulation and its fluctuations across the life course; for the longitudinal trajectories of AL see Tampubolon & Maharani [30]. Life course analyses require longitudinal analyses [39, 49].

## Conclusion

The life course shaping of allostatic load is at work in middle and high income nations, though in some high income ones the difference ceases to be significant. It appears that broader socioeconomic development has partly weakened the reach of early life adversity. This complexity in the shaping of health in later life is revealed for the first time thanks to the international harmonisation effort to collect blood biomarkers and the constructions of harmonised health risk factors. The WHO framework to implement the life course approach [40] can employ the methodology further afield. More importantly, future attribution of physiological dysregulation in later life can benefit from recognising the lifelong reach of early life adversity.

## Data Availability

All data produced are available online at http://www.icpsr.umich.edu/icpsrweb/ICPSR/studies/34315, https://www.share-project.org, https://www.elsa-project.ac.uk/, http://charls.pku.edu.cn/en/, https://www.rand.org/well-being/social-and-behavioral-policy/data/FLS/IFLS.html
https://tilda.tcd.ie/

https://www.share-project.org

http://charls.pku.edu.cn/en/

https://www.rand.org/well-being/social-and-behavioral-policy/data/FLS/IFLS.html

https://tilda.tcd.ie/

https://www.elsa-project.ac.uk/

## Supporting information

**S1 Table. Biomarker coverage**.

**S2 Table. Harmonised childhood items. S3 Table. Selection patterns**.

**S4 Table. Complete regression results**

## Acknowledgments

The World Health Organization (WHO) acknowledges financial support from Velux Stiftung, Zurich, including support to conduct research and advance metrics and evidence on healthy ageing across all stages of the life course. Disclaimer: VD, MC, CMJ, HYL, and RS are consultants or staff of the WHO; GT is the lead of WHO expert group (B3) on retrospective life course and healthy ageing. The authors alone are responsible for the views expressed in this publication and they do not necessarily represent the decisions, policy, or views of the WHO.

We thank the participants in 17 countries around the world for providing information, time and in many visits blood biomarkers too, as well as the generous funding bodies over many decades. HRS: The HRS (Health and Retirement Study) is sponsored by the National Institute on Aging (grant number NIA U01AG009740) and is conducted by the University of Michigan. ELSA: The English Longitudinal Study of Ageing was developed by a team of researchers based at University College London, NatCen Social Research, the Institute for Fiscal Studies, the University of Manchester and the University of East Anglia. The data were collected by NatCen Social Research. The funding is currently provided by the National Institute on Aging in the US (grant numbers: 2RO1AG7644 and 2RO1AG017644-01A1), and a consortium of UK government departments coordinated by the National Institute for Health Research. SHARE: The SHARE data collection has been funded by the European Commission, DG RTD through FP5 (QLK6-CT-2001-00360), FP6 (SHARE-I3: RII-CT-2006-062193, COMPARE: CIT5-CT-2005-028857, SHARELIFE: CIT4-CT-2006-028812), FP7 (SHARE-PREP: GA N°211909, SHARE-LEAP: GA N°227822, SHARE M4: GA N°261982, DASISH: GA N°283646) and Horizon 2020 (SHARE-DEV3: GA N°676536, SHARE-COHESION: GA N°870628, SERISS: GA N°654221, SSHOC: GA N°823782, SHARE-COVID19: GAN°101015924) and by DG Employment, Social Affairs & Inclusion through VS 2015/0195, VS 2016/0135, VS 2018/0285, VS 2019/0332, and VS 2020/0313. Additional funding from the German Ministry of Education and Research, the Max Planck Society for the Advancement of Science, the U.S. National Institute on Aging (U01 AG09740-13S2, P01 AG005842, P01 AG08291, P30 AG12815, R21 AG025169, Y1-AG-4553-01, IAG BSR06-11, OGHA 04-064, HHSN271201300071C, RAG052527A) and from various national funding sources (see www.share-project.org). TILDA: this research uses data from The Irish Longitudinal Study on Ageing, accessed via ISSDA (Trinity College Dublin, see https://idstilda.tcd.ie/data/access/) or ICPSR (University of Michigan http://www.icpsr.umich.edu/icpsrweb/ICPSR/studies/34315). CHARLS: the CHARLS research and field team coordinated by Peking University (http://charls.pku.edu.cn/en/). IFLS: the RAND Corporation, University of Indonesia, University of Gadjah Mada, SurveyMETER Yogyakarta (https://www.rand.org/well-being/social-and-behavioral-policy/data/FLS/IFLS.html).

## Notes

### Competing Interest Statement

The authors have declared no competing interest.

### Author Declarations

The study used ONLY openly available human data that were originally located at: http://www.icpsr.umich.edu/icpsrweb/ICPSR/studies/34315, https://www.share-project.org, https://www.elsa-project.ac.uk/, http://charls.pku.edu.cn/en/, https://www.rand.org/well-being/social-and-behavioral-policy/data/FLS/IFLS.html https://tilda.tcd.ie/

