## Supplemental Tables 1 - 4 for "Life course shaping of allostatic load across 17 countries: Evidence from England, the U.S., Europe, China and Indonesia"

### Supplementary Information

Supplement 1 Table. Summary of biomarkers for AL in HRS-family of ageing studies

| **Biomarkers** | **HRS** | **ELSA** | **TILDA** | **SHARE** | **CHARLS** | **IFLS** |
|---|---|---|---|---|---|---|
| Diastolic, systolic reading | ✓ | ✓ | ✓ | | ✓ | ✓ |
| Pulse wave velocity | | | | | | ✓ |
| Forced exp. volume | | | | ✓ | | |
| Cystatin C | ✓ | ✓ | ✓ | ✓ | ✓ | ✓ |
| C-reactive protein | ✓ | ✓ | ✓ | ✓ | ✓ | ✓ |
| Fibrinogen | | ✓ | | | | |
| Haemoglobin A1c | ✓ | ✓ | ✓ | ✓ | ✓ | ✓ |
| Haemoglobin (Hb) | | | | | | ✓ |
| High-density lipoprotein/ total cholesterol ratio | ✓ | ✓ | ✓ | ✓ | ✓ | |
| Triglycerides | | ✓ | ✓ | ✓ | ✓ | |
| Waist circumference | ✓ | | | | ✓ | |
| Waist-hip ratio | | | | | | ✓ |
| BMI | ✓ | ✓ | ✓ | ✓ | ✓ | ✓ |

Supplement 2 Table. Summary of included items for childhood poverty construct

| **HRS** | **ELSA** | **TILDA** | **SHARE** | **CHARLS** | **IFLS** |
|---|---|---|---|---|---|
| Number of rooms | Number of rooms | Number of rooms | Number of rooms | | Number of rooms |
| Number of people | Number of people | Number of people | | | Number of people |
| Number of books | Number of books | Number of books | | | Number of books |
| | Physical facilities: indoor toilet, hot and cold running water, central heating, fixed bath | Physical facilities: indoor toilet, hot and cold running water, central heating, fixed bath | Physical facilities: indoor toilet, hot and cold running water, central heating, fixed bath. | | Physical facilities: piped water, outdoor toilet with septic tank. |
| Ever experienced: being financially poor, had to move or be helped because of financial difficulty, had to live with grandparents, father unemployed | | Financial hardship during childhood | | Financial hardship during childhood, family had to move because of risk of starving, family member starved to death during the Great Leap Forward, neighbourhood unclean or unsafe, neighbours unwilling to help, father unemployed, parent illiterate, father unemployed. | |

Supplement 3 Table. Missing data or non-response patterns across analytic vs declined sample in HRS, ELSA, SHARE, TILDA, CHARLS and IFLS

| | Variable | Declined Sample[a] | Analytic Sample[b] |
|---|---|---|---|
| **HRS**[c] | Female | 56.04% | 57.52% |
| | Male | 43.96% | 42.48% |
| | | $\chi^2 = 2.382$, $p = 0.123$ | |
| | Mean age | 66.84 year | 67.00 year |
| | | $p = 0.476$ | |
| **ELSA** | Female | 54.49% | 55.49% |
| | Male | 45.51% | 44.51% |
| | | $\chi^2 = 0.982$, $p = 0.335$ | |
| | Mean age | 68.85 year | 71.82 year |
| | | $p = 0.000$ | |
| **SHARE** | Female | 55.52% | 58.13% |
| | Male | 44.48% | 41.87% |
| | | $\chi^2 = 24.243$, $p = 0.000$ | |
| | Mean age | 67.16 year | 67.72 year |
| | | $p = 0.000$ | |
| **TILDA** | Female | 55.55% | 53.69% |
| | Male | 44.45% | 46.31% |
| | | $\chi^2 = 9.694$, $p = 0.002$ | |
| | Mean age | 62.97 year | 61.74 year |
| | | $p = 0.000$ | |
| **CHARLS** | Female | 51.78% | 51.55% |
| | Male | 48.22% | 48.45% |
| | | $\chi^2 = 0.135$, $p = 0.173$ | |
| | Mean age[d] | 58.49 year | 62.85 year |
| | | $p = 0.000$ | |
| **IFLS** | Female | 52.89% | 61.71% |
| | Male | 47.11% | 38.29% |
| | | $\chi^2 = 9.570$, $p = 0.002$ | |
| | Mean age[e] | 58.24 year | 57.93 year |
| | | $p = 0.000$ | |

Note: [a] Main survey wave for each cohort. [b] Participants excluded if any AL component is missing. [c] Only White Americans. [d] Aged 50 or older. [e] Aged 45 or older.

Supplement 4 Table. Linear regression coefficients and confidence interval (95%) of allostatic load in the US (HRS), England (ELSA), Europe (SHARE), Ireland (TILDA), China (CHARLS) and Indonesia (IFLS)

| | **HRS** | | | **ELSA** | | | **SHARE** | |
|---|---|---|---|---|---|---|---|---|
| **N. Obs** | 2427 | 2427 | 2427 | 2038 | 2038 | 2038 | 8184 | 8184 |
| Childhood poor | .037 | .026 | -.037 | .027 | .018 | -.002 | .007 | -.001 |
| | (-.039, .113) | (-.050, .101) | (-.114, .040) | (-0.018, 0.072) | (-.028, .063) | (-.050, .046) | (-.014, .028) | (-.023, .020) |
| Age | | .005 | .006*** | | .003 | .002* | | .002 |
| | | (.003, .006) | (.004, .008) | | (.001, .005) | (.000, .005) | | (.001, .003) |
| Sex, female | | -.066*** | -.080*** | | .024 | -.002 | | -.152*** |
| | | (-.101, -.032) | (-.114, -.045) | | (-.012, .060) | (-.040, .036) | | (-.168, -.136) |
| Some college | | | -.093* | | | -.050 | | |
| | | | (-.165, -.021) | | | (-.113, .012) | | |
| Rural (non-urban) | | | -.002 | | | -.015 | | |
| | | | (-.036, .032) | | | (-.055, .026) | | |
| Unmarried/Single | | | .014 | | | .023 | | |
| | | | (-.025, .052) | | | (-.758, .804) | | |
| Poorest quartile[a] | | | .198*** | | | .128*** | | |
| | | | (.149, .247) | | | (.072, .183) | | |
| Constant | -.071*** | -.325*** | -.407*** | -.003 | -.224** | -.211 | .022*** | -.010 |
| | (-.088, -.053) | (-.448, -.202) | (-.570, -.245) | (-.023, .017) | (-.379, -.069) | (-1.021, .599) | (.013, .0308) | (-.064, .043) |

| | **TILDA** | | | **CHARLS** | | | **IFLS** | |
|---|---|---|---|---|---|---|---|---|
| **N. Obs** | 4079 | 4079 | 4079 | 5220 | 5220 | 5220 | 1243 | 1243 |
| Childhood poor | .071*** | .051* | .038 | .086*** | .037 | .035* | .132** | .140** |
| | (.040, .101) | (.022, .081) | (.007, .068) | (.053, .118) | (.003, .070) | (.001, .070) | (.053, .211) | (.059, .221) |
| Age | | .011*** | .011*** | | .008*** | .007*** | | .001 |
| | | (.010, .013) | (.009, .012) | | (.006, .009) | (.006, .008) | | (-.002, .005) |
| Sex, female | | -.125*** | -.125*** | | -.028** | -.031** | | -.124*** |
| | | (-.149, -.101) | (-.149, -.101) | | (-.049, -.008) | (-.053, -.010) | | (-.181, -.067) |
| Some college | | | -.060** | | | .138** | | |
| | | | (-.094, -.026) | | | (.060, .216) | | |
| Rural (non-urban) | | | .000 | | | -.056*** | | |
| | | | (-.024, .024) | | | (-.082, -.029) | | |
| Unmarried/Single | | | .004 | | | .013 | | |
| | | | (-.024, .032) | | | (-.060, .087) | | |
| Poorest quartile[a] | | | .032* | | | .023 | | |
| | | | (.001, .064) | | | (-.009, .054) | | |
| Constant | -.020** | -.637*** | -.605*** | .019** | -.443*** | -.383*** | .110*** | .107 |
| | (-.033, -.006) | (-.729, -.546) | (-.715, -.496) | (.008, .030) | (-.518, -.367) | (-.475, -.292) | (.080, .142) | (-.071, .286) |

Note: Significant level: *5%, **1%, ***0.1%. [a] current non-housing financial asset.
